# Rapid SARS-CoV-2 testing in primary material based on a novel multiplex LAMP assay

**DOI:** 10.1101/2020.06.18.20130377

**Authors:** Bernhard Schermer, Francesca Fabretti, Maximilian Damagnez, Veronica Di Cristanziano, Eva Heger, Sita Arjune, Nathan A. Tanner, Thomas Imhof, Manuel Koch, Alim Ladha, Julia Joung, Jonathan S. Gootenberg, Omar O. Abudayyeh, Volker Burst, Feng Zhang, Florian Klein, Thomas Benzing, Roman-U. Müller

## Abstract

**Background:** Rapid and extensive testing of large parts of the population and specific subgroups is crucial for proper management of severe acute respiratory syndrome coronavirus 2 (SARS-CoV-2) infections and decision-making in times of a pandemic outbreak. However, point-of-care (POC) testing in places such as emergency units, outpatient clinics, airport security points or the entrance of any public building is a major challenge. The need for thermal cycling and nucleic acid isolation hampers the use of standard PCR-based methods for this purpose.

**Methods:** To avoid these obstacles, we tested PCR-independent methods for the detection of SARS-CoV-2 RNA from primary material (nasopharyngeal swabs) including loop-mediated isothermal amplification (LAMP) and specific high-sensitivity enzymatic reporter unlocking (SHERLOCK).

**Results:** Whilst specificity of standard LAMP assays appears to be satisfactory, sensitivity does not reach the current gold-standard quantitative real-time polymerase chain reaction (qPCR) assays yet. We describe a novel multiplexed LAMP approach and validate its sensitivity on primary samples. This approach allows for fast and reliable identification of infected individuals. Primer optimization and multiplexing helps to increase sensitivity significantly. In addition, we directly compare and combine our novel LAMP assays with SHERLOCK.

**Conclusion:** In summary, this approach reveals one-step multiplexed LAMP assays as a prime-option for the development of easy and cheap POC test kits.

## Introduction

The recent pandemic of SARS-CoV-2 is a major challenge for healthcare systems worldwide. Lacking an effective and approved vaccine, reliable screening of samples from nasopharyngeal swabs for viral RNA is a fundamental pillar for effective disease control. Governmental measures (e.g. shutdown of social life) largely depend on such data to allow for a rapid adaptation to changes in epidemiologic indicators. Furthermore, isolation and quarantine of infected and exposed individuals are key for effective outbreak control. Here, the prompt establishment of a diagnostic real-time PCR based testing *(1)* has been instrumental already in the early phases of the pandemic. Such qPCR-based tests are extremely powerful due to their high sensitivity and specificity. However, these assays require specific lab equipment and expertise which is typically not widely available directly at the point-of-care making transportation to a specialized facility necessary. Furthermore, the material required for the different steps involved may become subject to shortages during a pandemic making alternative approaches an important goal. Even though RNA isolation and subsequent qPCR is performed in only a few hours, actual turnaround times from sample collection to diagnostic test results are often much longer. Decision-making in both healthcare facilities and other spheres of public life would be facilitated enormously by direct testing on site with short turn-around times. Such an approach could also limit the need for quarantine, allow healthcare workers after exposure to continue their work upon a daily negative swab and avoid shortages in systemically relevant personnel. Recently, a number of PCR-independent methods have been proposed for this purpose including isothermal amplification (RPA, LAMP *(2–10)*) and their combination with genome editing tools such as Cas12a *(11)* or Cas13a *(12, 13)* for improved performance. The majority of these studies, however, have not been carried out on direct primary material but on RNA isolated from patient samples or generated in vitro *(3–9, 11)*. In the study at hand, we use previously described LAMP and SHERLOCK assays on both isolated RNA and primary material from patients *(9, 12)*. In addition, we describe a newly designed multiplexed LAMP assay targeting Orf3a and Orf7a of SARS-CoV-2. Orf3a and Orf7a have been selected since these targets have not been used for any available diagnostic qPCR assay, to avoid the risk of amplicon cross-contamination between different types of assays.

## Methods

### Samples in universal transfer medium and isolated RNA

Nasopharyngeal swabs were taken from symptomatic patients presenting at University Hospital of Cologne from March to April 2020. Swabs were directly transferred in 1-3 ml of universal transfer medium (UTM) or PBS. For diagnostic qPCR, RNA was extracted from 500 µl UTM / PBS of the swab samples using the automated MagNA Pure 96 system (Roche). 0.5 to 1.9 µl isolated RNA or 0.5 to 1.9 µl UTM/PBS was used for subsequent LAMP/Sherlock assays. Diagnostic qPCR was performed using a RealStar SARS-CoV-2 RT-PCR kit 1.0 targeting E and S gene (Altona Diagnostics) on a LightCycler 480 (Roche).

### Sample inactivation for LAMP and Sherlock

For assays from direct material 10 µl of UTM/PBS were incubated at 98°C for 15 minutes in a PCR cycler with heated lid placed inside a safety cabinet. Treatment of 15 min at 92°C has been shown to inactivate SARS-CoV-2 *(14)* and 98°C was found to be preferable for downstream RNA-based applications.

### LAMP primer design

For Orf1a and Gene N we used previously described primer sets *(9)*. Additional primer sets were designed for Orf3a, Orf7a and the M gene using the primerexplorer V5 tool (http://primerexplorer.jp/e/). All primers were ordered from IDT, purified with standard desalting, as PAGE purification did not improve the performance of the assays (data not shown). All primer sequences are listed in supplementary table 1.

### LAMP assay

All LAMP reactions were assembled on ice. In brief, each 20 µl reaction contained 10 µl WarmStart Colorimetric LAMP mix (NEB), 2 µl primer mix (F3/B3 2 µM each; FIP/BIP 16 µM each; LF/LB 4 µM each), 40 mM guanidine hydrochloride (from a 4 M stock solution, pH8), DNase/RNase free water and 0,5 µl sample. For multiplexed LAMP the additional primer mix replaced 2 µl of DNase/RNAse free water. Initially, the assays were performed without guanidine hydrochloride at 65°C for 40 minutes, while the multiplexed reaction was done at 60°C for 40 to 50 minutes, as indicated in the figures. Each assay was performed including several negative controls. Positive reactions were identified due to a clear change in color from pink/red to orange/yellow. In two cases out of almost 200, the mere addition of 0.5 µl sample to the reaction resulted in a color change. These samples are not included in the data shown.

### SHERLOCK Assay with RPA

The two step SHERLOCK assay for Orf1a and S-gene was performed according to the protocol established in Feng Zhang’s lab at MIT (https://zlab.bio/s/COVID-19-detection-v20200321.pdf) (12, 13). In brief, an RPA reaction was set up using 5.9 µl RPA mix (Twist AMP), 0.2 µl Protoscript RT (NEB), 1µl RPA primer mix (10 µM each), 1.9 µl sample (isolated RNA or UTM/PBS) and 0.5 µl magnesium acetate (280 mM stock solution). The reaction was mixed, spun down and incubated 42°C for 25 min. The Cas13a-based detection step was performed in a 20 µl reaction as follows: 2 µl TRIS-HCl buffer (400 mM, pH 7.4), 9.6 µl DNase/RNase free water, 2 µl LwaCas13a (corresponding to 120 ng of protein), 1 µl crRNA (10 ng/µl), 1 µl Lateral-Flow-Reporter (20 µM), 1 µl SUPERase In RNase Inhibitor (Thermofisher Scientific), 0.6 µl T7 Polymerase (Lucigen), 0.8 µl rNTPs (25 µM each; NEB), 1 µl MgCl2 (120 mM stock solution) and 1 µl of previous RPA reaction. The reaction was mixed and spun down, then incubated at 37°C for 30 min. Next, the reaction was diluted with 80 µl of HybriDetect Buffer (Milenia), mixed, and then a HybriDetect dipstick (Milenia) was placed in the reaction tube and incubated at RT. Results were visible within 5 minutes.

### Cas13a combined with LAMP

To combine the LAMP amplification with Cas13a detection we added a T7 promoter in the loop region of the FIP primers for the LAMP assay (suppl. table 1). After the LAMP reaction at 65°C for 40 min, 0.5 µl were used in the Cas13a detection assay assembled as described above, using a specific crRNA targeting the LAMP Gene N amplicon.

## Results

To allow for the comparison of different nucleic acid detection methods for SARS-CoV-2 we collected redundant material from nasopharyngeal swabs obtained for qPCR testing in clinical routine due to suspected COVID-19. 171 samples were selected randomly from samples collected in the period from March to April 2020. The cohort included individuals between the age of 1 month and 88 years with a close to equal distribution of men and women (Suppl. Table 2).

### SHERLOCK and LAMP on isolated RNAs

We first tested two recently described assays for SARS-CoV-2 detection on isolated RNA from patient samples. Specifically, we performed colorimetric LAMP assays using primers targeting Orf1a and Gene-N *(9)* and two-step SHERLOCK assays combined with lateral flow detection *(12, 13)* targeting Orf1a and S gene (Suppl. Fig. 1, A-C). Even though both assays worked well on these samples, they failed to detect the virus in specimen that were positive in diagnostic qPCR at cycle threshold (Ct) values > 30 for E and S gene (Fig. S1). Moreover, LAMP appeared to be slightly more sensitive and specific (Fig. S1C).

**Fig. 1:**
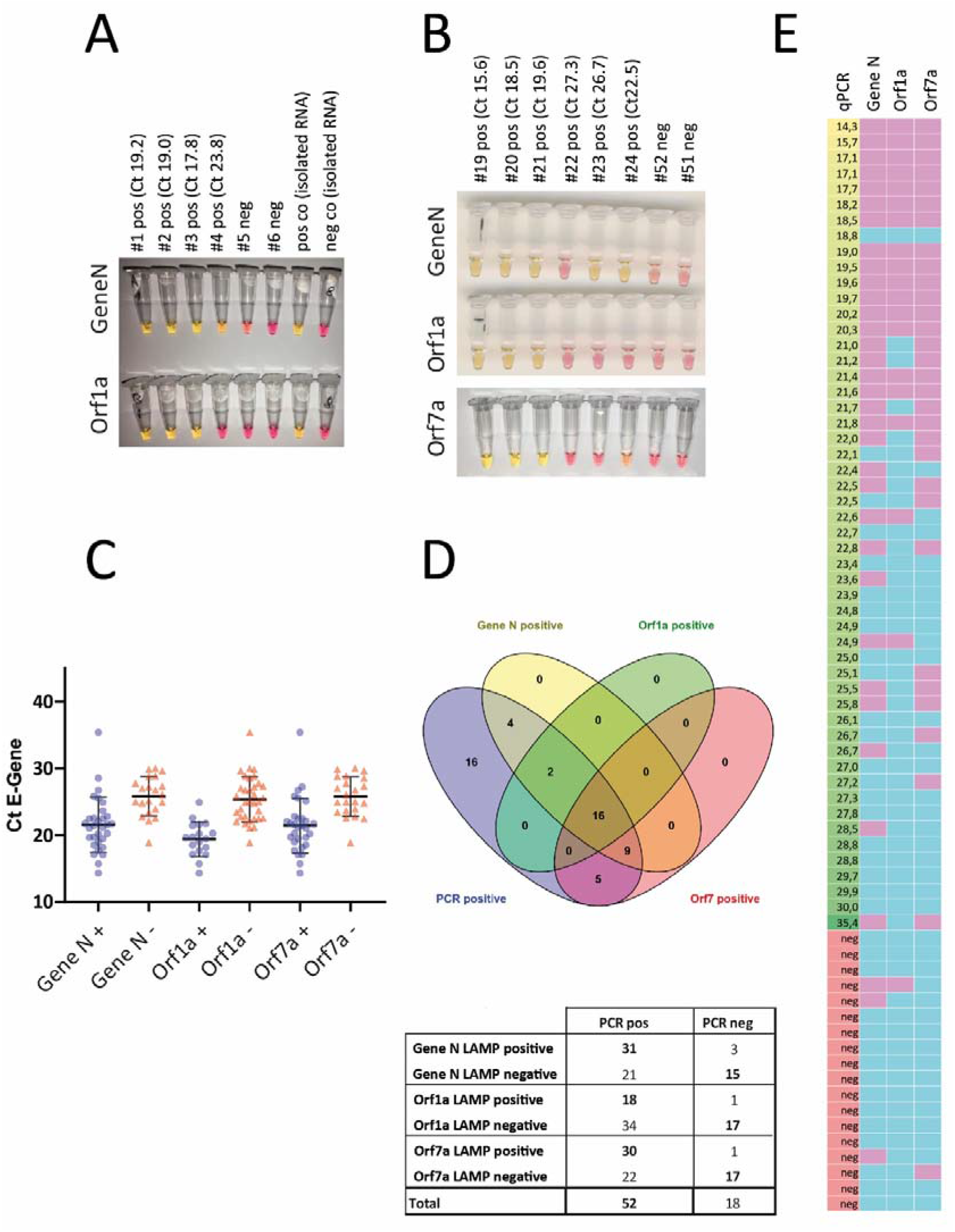
Detection of viral RNA with LAMP. A. Representative pictures of a LAMP assay. UTM samples from 6 patients, 4 of which tested positively for SARS-CoV-2 in diagnostic qPCR were analyzed by LAMP targeting Gene N and Orf1a. Isolated RNA from swabs served as positive and negative control. B. 70 samples were analyzed with three different LAMP assays targeting Gene N, Orf1a and Orf7a. Representative results from additional assays. C. The individual value plot shows of Ct values of the diagnostic qPCR for the E gene of positive and negative LAMP assays. D. Total number of positive and negative LAMP assays from patient samples tested positive or negative for SARS-CoV-2. E. Heat map summarizing the results of three different LAMP assays on all 70 samples (left column: Ct value for E gene from diagnostic PCR in ascending order; other columns: pink indicating positive, blue indicating negative result. Grey: not performed)

### LAMP assays on primary material from nasopharyngeal swabs

Due to limited availability of the RPA reagents required for the first step of SHERLOCK at that time paired with the results described above, we focused our efforts on validation of the LAMP assays. Since detection worked on isolated RNA, we went on to test this approach using primary material (i.e. transport medium from nasopharyngeal swabs without RNA isolation) Strikingly, both LAMP assays performed well on these specimens. Using as little as 0.5 µl UTM as input for each reaction to avoid inhibitory effects of the medium or of tissue contaminants on the reaction, detection worked in samples tested positive by qPCR (see Ct as reference) (Fig. 1A).

### Establishment of primer sets for novel target genes in LAMP assays

In parallel, 5 additional sets of LAMP primers targeting additional genes of the SARS-CoV-2 genome (NC_045512) were designed, tested and validated. Here, we preferred genes that are not target of any standard diagnostic qPCR assay, to minimize and avoid any possible interference of such point of care assays with other routine diagnostic pipelines. Among these new primer sets, the oligos targeting Orf7a showed the highest sensitivity and specificity in several tests on diluted isolated RNA (data not shown) and was thus selected for further testing in primary material. In this second set of experiments we screened a total of 70 samples, 52 of which had been tested positive for SARS-CoV-2 by diagnostic qPCR. Fig. 1B displays representative results of the LAMP assays targeting Gene N, Orf1a and Orf7a. Assays targeting Gene N and Orf7a were more sensitive than the one targeting Orf1a as indicated by the mean Ct value of LAMP-positive samples (Fig. 1C), by the total number of samples that were correctly identified (Fig. 1D/Fig. 1E). 3 out of 70 specimens turned out positive for Gene N and 1 out of 70 for Orf1a and Orf7 respectively, that were negative by qPCR.

### Increasing the sensitivity of LAMP

In order to increase the sensitivity of LAMP assays to a comparable level with qPCR we performed a “two-step” LAMP reaction, either using a small amount (0.2 to 0.5 µl) of a first LAMP reaction as template for a second one or by refreshing the first reaction with new enzymes and dNTPs. All of these attempts resulted consistently in negative controls (water, empty UTM or samples from negative patients) turning positive, either because of unspecific amplification or because of the necessary re-opening of the reaction tubes after the first amplification step. The use of mineral oil on top of every reaction – performed to avoid cross-contamination – did not improve these results (data not shown). In an additional set of experiments, we combined the LAMP assays targeting Gene N with LwaCas13a mediated detection. To this end, we added a T7 promoter in the loop region of the FIP-primer and designed a crRNA targeting the amplicon of the LAMP reaction. The addition of the T7 promotor did not affect the efficiency of the LAMP reaction (Suppl. Fig. 2A) and a positive LwaCas13a mediated detection is indicated by the upper band in the lateral flow assay (Suppl. Fig. 2B). While this combination of LAMP and LwaCas13a provides an additional proof of specificity as compared to the LAMP reaction alone, sensitivity was not increased. An additional possibility to improve sensitivity of LAMP is multiplexing, using primer mixes for different genes of the viral genome in the same reaction. After testing different combinations of these primer sets, we observed superior performance of LAMP assays containing primer sets amplifying Orf7a and Orf3a combined with a lower reaction temperature (Suppl. Fig. 3).

### Multiplex LAMP assay on clinical samples

To increase accessibility of RNA and efficiently inactivate the virus we incubated UTM from swabs at 98°C for 15 min. Treatment at 92°C for 15 minutes has been demonstrated to efficiently inactivate SARS-CoV-2 *(14)*, while 98°C appeared a good choice for downstream RNA applications in case of SARS-CoV-2 *(15)*. After incubation at 98 °C, some of the UTM samples showed a gel-like consistency. In these cases, pipetting about 10 times up and down with a P20 pipette allowed for further accurate pipetting. Again, we only used 0.5 µl UTM for each reaction. In addition, we added guanidine hydrochloride as a classical RNAse inhibitor and LAMP enhancer*(16)* to the multiplexed reaction at 60°C. These modifications in sample preparation combined with our new multiplexed assay were used to analyze a set of 102 clinical samples consisting of 74 SARS-CoV-2 positive and 28 negative swabs (Fig. 2). 54 SARS-CoV-2 positive samples with Ct values up to 38 were positive in the LAMP assay (Fig. 2 B), while still 20 out of 74 qPCR positive samples were not detected in our assay on direct material. However, the vast majority of samples up to a Ct of 30 were correctly identified (94%; 45 out of 48). Meanwhile, RPA reagents arrived and we performed the two step SHERLOCK assays on some of the very same direct samples. However, sensitivity was much lower with a cut-off threshold of 21 cycles (Fig. 2D; representative assays shown in Suppl. Fig. 4). In summary, our multiplex LAMP protocol is a simple and sensitive way to detect SARS-CoV-2 RNA from clinical samples.

**Fig. 2:**
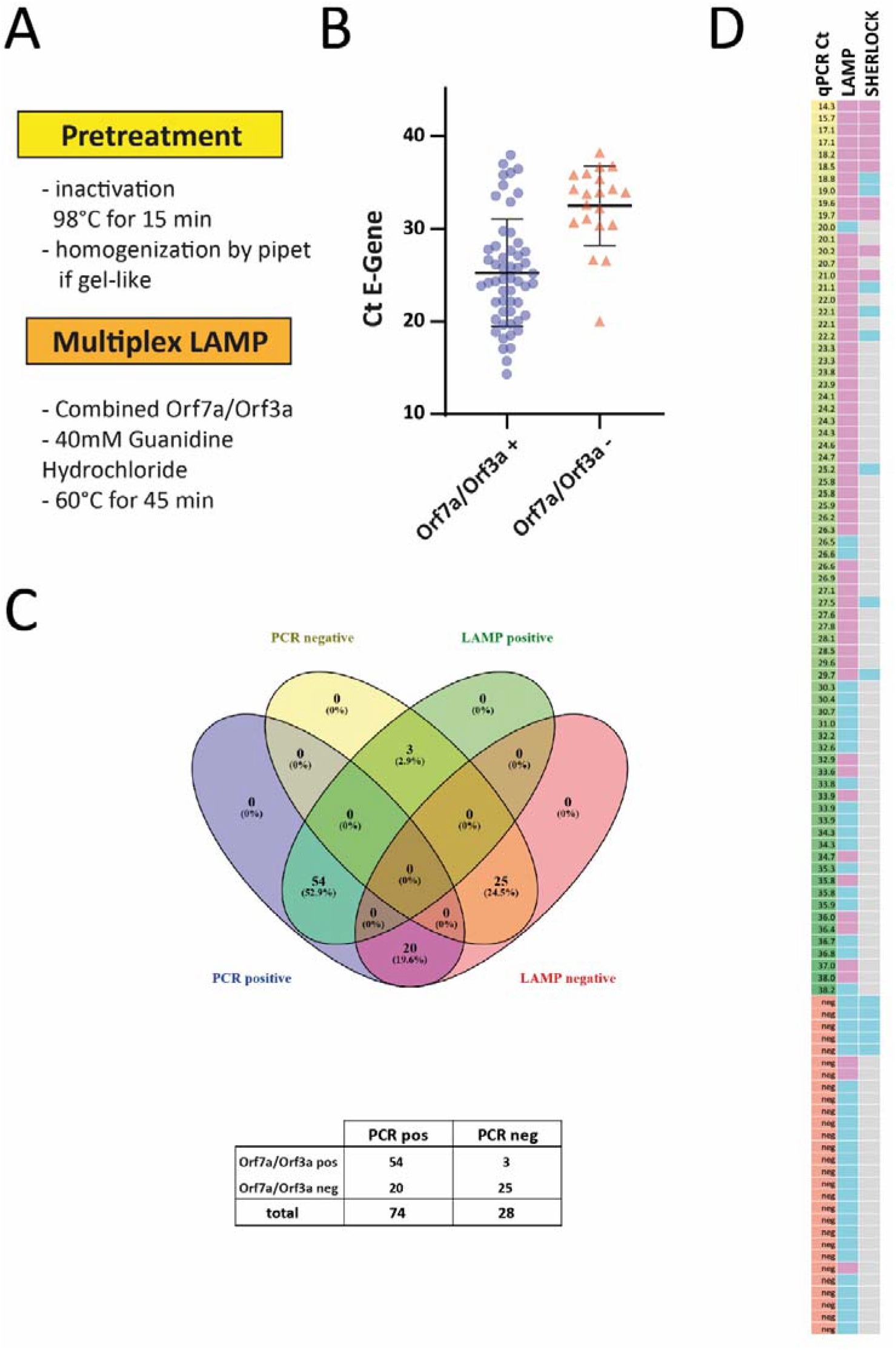
Detection of viral RNA with multiplexed LAMP. A. Workflow of the optimized multiplexed LAMP protocol. B. 104 samples were analyzed with the multiplexed LAMP assay targeting Orf7a and Orf3a. The individual value plot shows Ct values of the diagnostic qPCR for the E gene of positive and negative LAMP assays. C. Total number of positive and negative multiplexed LAMP assays from patient samples tested positive or negative for SARS-CoV-2. D. Heat map summarizing the results (left column: Ct value for E gene from diagnostic PCR in ascending order; middle and right column: pink indicating positive, blue indicating negative result. Grey: not performed).

## Discussion

Based on our data we conclude, that multiplexing primers in LAMP reactions is a highly promising way to further increase sensitivity of these assays and to quickly develop a rapid POC test. Currently, a test based on our multiplexed LAMP assay would – in contrast to a good specificity - most likely miss to identify those infected patients with very low amounts of viral RNA in the nose or throat and would not yet reach the sensitivity of the gold-standard qPCR assays. However, the LAMP approach comes with several clear-cut advantages. In contrast to classical qPCR, it can be used in primary material without the need for RNA isolation and yields results rapidly. Furthermore, only little equipment is required (thermoblock) and could easily be made available in e.g. emergency rooms. The estimated costs for reagents are below 1.50 € per reaction with the advantage – compared to qPCR – that additional expenses for nucleic acid isolation are not required. Regarding the target setting of primary screening, one could hypothesize, that identifying patients with high viral loads would detect those individuals that are highly infectious. Consequently, in this setting, even a POC test with a lower sensitivity as compared to qPCR would be useful and could be backed up by a combination with qPCR the results of which get available later. Currently, a relation between high viral RNA in swabs and the infectivity of a patient has not yet been definitively established and detection of viral RNA is not equivalent to detection of infectious virus. One retrospective study, however, suggested low infectivity of patients with Ct-values >24 in E gene qPCR from swabs since the authors did not observe viral growth in exposed Vero cells *(17)*. Besides, a statement paper of the National Centre for Infectious Diseases and the Chapter of Infectious Disease Physicians (Singapore) refers to a study with 73 COVID-19 patients where a Ct >30 was found to be the threshold of infectivity *(18)*. Nonetheless, the ultimate goal would be a POC test that reaches the sensitivity of qPCR and can completely replace the current approach where favorable. Studies in the near future will clarify, whether patients with high viral RNA loads are indeed the most contagious individuals. Besides, this knowledge will also be of greatest importance for the actual clinical interpretation of qPCR results in the future. By now, it has been shown that the viral load is already high before and maybe highest at onset of symptoms *(19)* and at the time point of presentation to the clinic followed by a steady decline *(20)*. Consequently, when focusing the screening on pre-symptomatic individuals, sensitivity of the LAMP assay may actually be higher than in the current cohort. Additionally, one thing to be kept in mind when directly comparing sensitivity between the different methods, is the fact that qPCR reaches this standard only in isolated RNA whilst the multiplex LAMP assay attains an optimized detection rate already in primary material. Regarding specificity, only few specimens turned out to be positive in the multiplex LAMP assay that were negative in qPCR. Since qPCR is the gold-standard this can be interpreted as a minor limitation in specificity. However, qPCR itself does not reach a sensitivity of 100% *(21)*. Consequently, it is not clear yet whether these individuals were truly negative or missed by the qPCR assay.

CRISPR/ Cas12 *(11)* or Cas13a *(13)* based assays are another promising way to detect RNA in a PCR-independent manner. Comparison of this approach with LAMP demonstrates a surprisingly high sensitivity even of the colorimetric one step LAMP assay. Very recently, a novel protocol for Cas13a - called STOP (‘SHERLOCK Testing in One Pot’) - has been described *(22)*. As this replaces the isothermal RPA reaction of the original SHERLOCK protocol by a LAMP reaction, the authors use a thermo-stable Cas13a enzyme to enable performing the entire reaction at the same temperature. Whilst being a very interesting approach, nonetheless, this comes with the difficulty the reaction tube has to be opened for the final lateral flow assay used for detection. In the real-world POC testing setting, this would require the establishment of a ‘pre-amp’ and ‘post-amp’ area to avoid cross-contamination, which may limit its use. Alternatively, lateral flow could be replaced by using a fluorescent probe together with an appropriate simple detection device. However, the highest Ct value resulting in a positive STOP assay is - at about 30 cycles - in a similar range compared to a recently described Cas12a-based method (DETECTR) *(22)*. The LAMP assay described in our study works without re-opening the test tube after amplification and provides detection at higher Ct values. On the other side, both SHERLOCK and DETECTR add an additional level of specificity to the detection due to the crRNA directing the Cas12/13 enzyme. Both SHERLOCK and DETECTR require a considerable number of pipetting steps. In contrast, the multiplexed LAMP assay demonstrates a similar or even higher sensitivity and requires only two simple pipetting steps at the POC: *(1)* taking a aliquot of the UTM for ‘boiling’ and *(2)* the addition of the 0.5 µl sample to the reaction tube, which could also be reached without a pipet by an inoculating loop. Reaction tubes can be prepared in anticipation elsewhere (e.g. in any nearby central facility) and stored for hours at 4°C. The preparation of these reaction tubes, is also done with only four simple pipetting steps (1. primer mix pre-diluted in water, 2. LAMP mix, 3. guanidine hydrochloride, → aliquot in 200 µl tubes). This could even be further simplified by adding guanidine to the primer mix. After amplification, the tube and the according controls are photo-documented and discarded. In summary, we are convinced that systematically combining and testing different multiplex LAMP primer sets on primary swab material is one of the most promising approaches to develop a powerful POC test.

## Data Availability

All data is available. All protocols will be provided.

## Author Contributions

RUM and BS initiated this project. BS designed and performed most experiments. BS and FF designed primers and crRNAs. VdC, MD, EH, VB and FK provided patient material and Ct values from diagnostic qPCR. SA collected and prepared clinical data from the cohorts. FZ, OOA, JSG, JJ, and AL provided plasmids, reagents (SHERLOCK test kit) and detailed instructions for the SHERLOCK detection. Cas13a subcloning, expression and purification was done by TI and MK. BS and FF prepared the figures. BS, FF and RUM wrote the manuscript, which was critically revised by VB, MK, TB, VdC, EH, NAT and FK.

## Acknowledgments

We thank Stefanie Keller for her outstanding technical support during very special and challenging times. Special thanks to the team of Benchling for revolutionizing the work with DNA/RNA sequences. Venn diagrams were generated using “Venny 2.1” by J.C. Oliveros (https://bioinfogp.cnb.csic.es/tools/venny/index.html, accessed in May 2020). This work was supported by the German Federal Ministry for Education and Research (01KI20184 to BS, VB, RUM und FK) and by the German Research Foundation [CRU329 to BS, RUM and TB].

## Non-standard abbreviations

COVID-19: coronavirus disease-2019
SARS-CoV-2: severe acute respiratory syndrome coronavirus 2
qPCR: quantitative real-time polymerase chain reaction
UTM: universal transport media
POC: point-of-care
LAMP: loop-mediated isothermal amplification
SHERLOCK: specific high-sensitivity enzymatic reporter unlocking
RPA: recombinase polymerase amplification
Ct: cycle threshold

## Figure legends

**Fig. S1:**
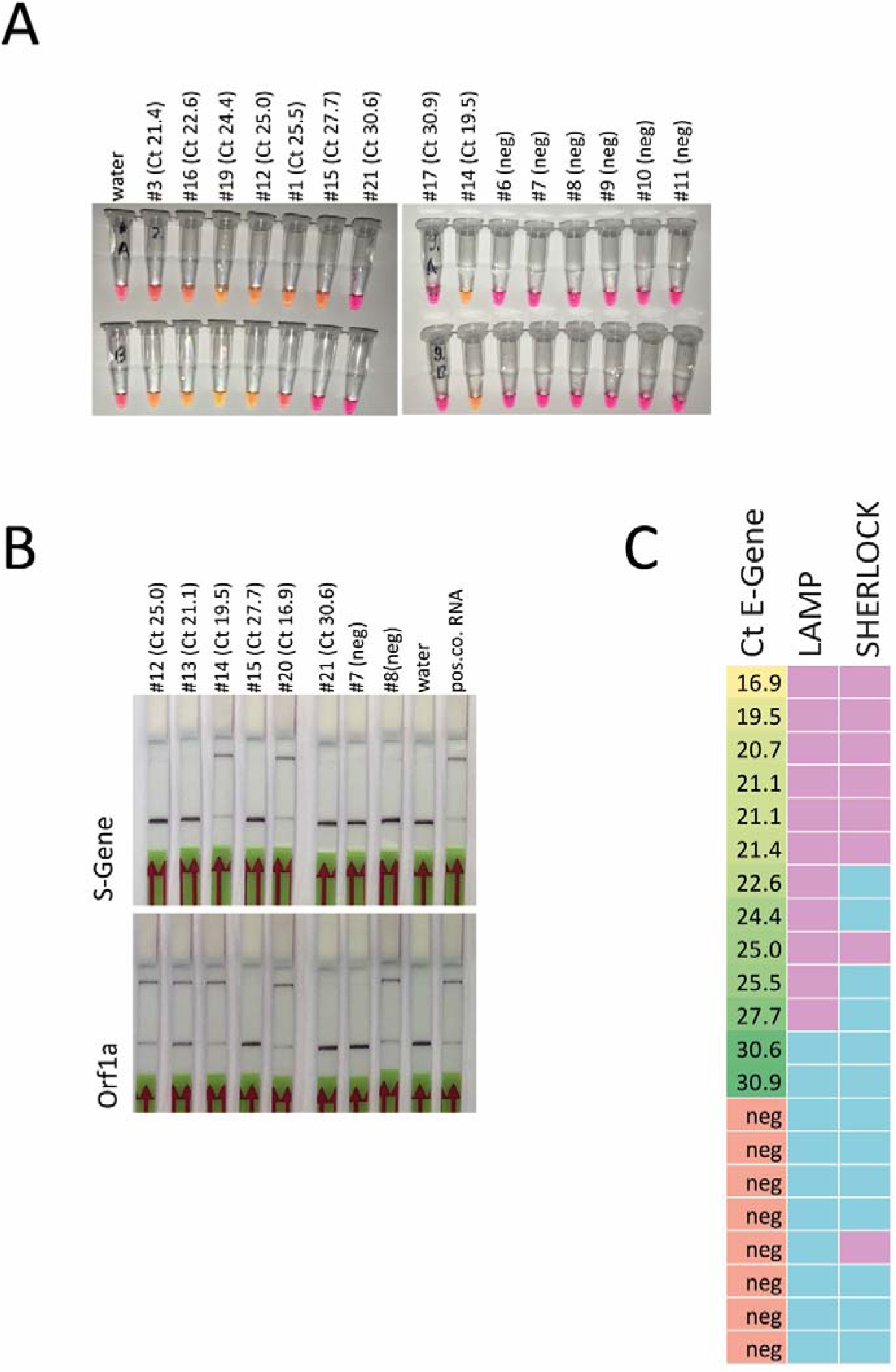
Detection of viral RNA with LAMP and SHERLOCK using isolated RNA from swabs. A. representative result from LAMP assay targeting Gene N and Orf1a. Shift from red/pink to yellow/orange indicates a positive result (Ct value of E gene from diagnostic qPCR). B. Sherlock assay for S gene and Orf1a. The upper band indicates a positive result while the lower band is a control (Ct value of E gene from diagnostic qPCR). C. Summary of the LAMP and Sherlock results on all available RNA samples (assay was regarded as positive with at least one positive result out of two LAMP or SHERLOCK assays; left column: Ct value for E gene from diagnostic PCR in ascending order; green: positive result, blue negative result).

**Fig. S2:**
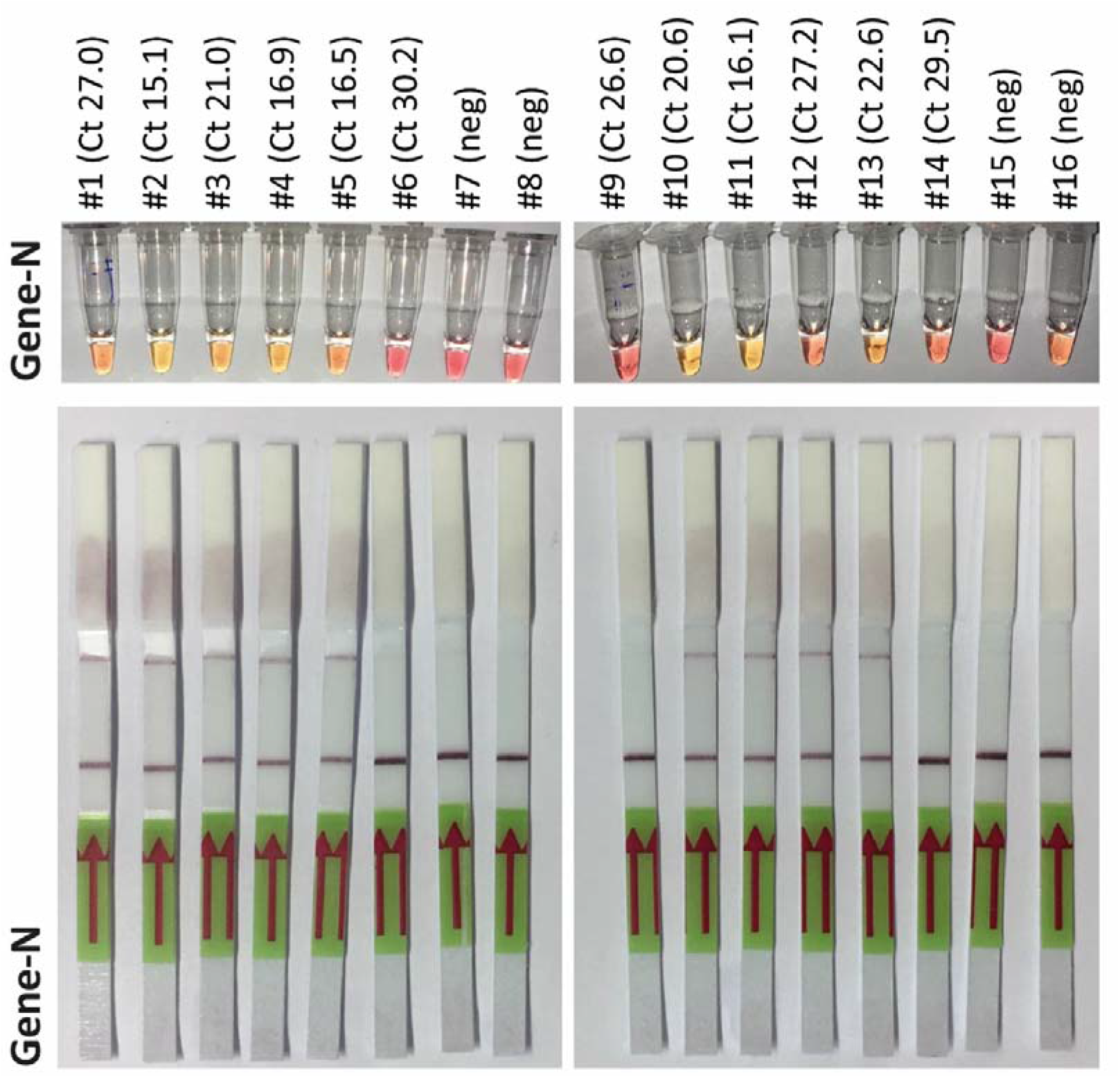
Detection of viral RNA with LAMP and LwaCas13a. A. Results from the colorimetric LAMP assay using 0,5 µl sample (UTM) with primers containing a T7 promotor in the loop region. B. 1 µl of the LAMP reaction from A was entered in the Cas13a recognition reaction followed by lateral flow assay. Upper band indicates positive recognition of the Gene N target sequence.

**Fig. S3:**
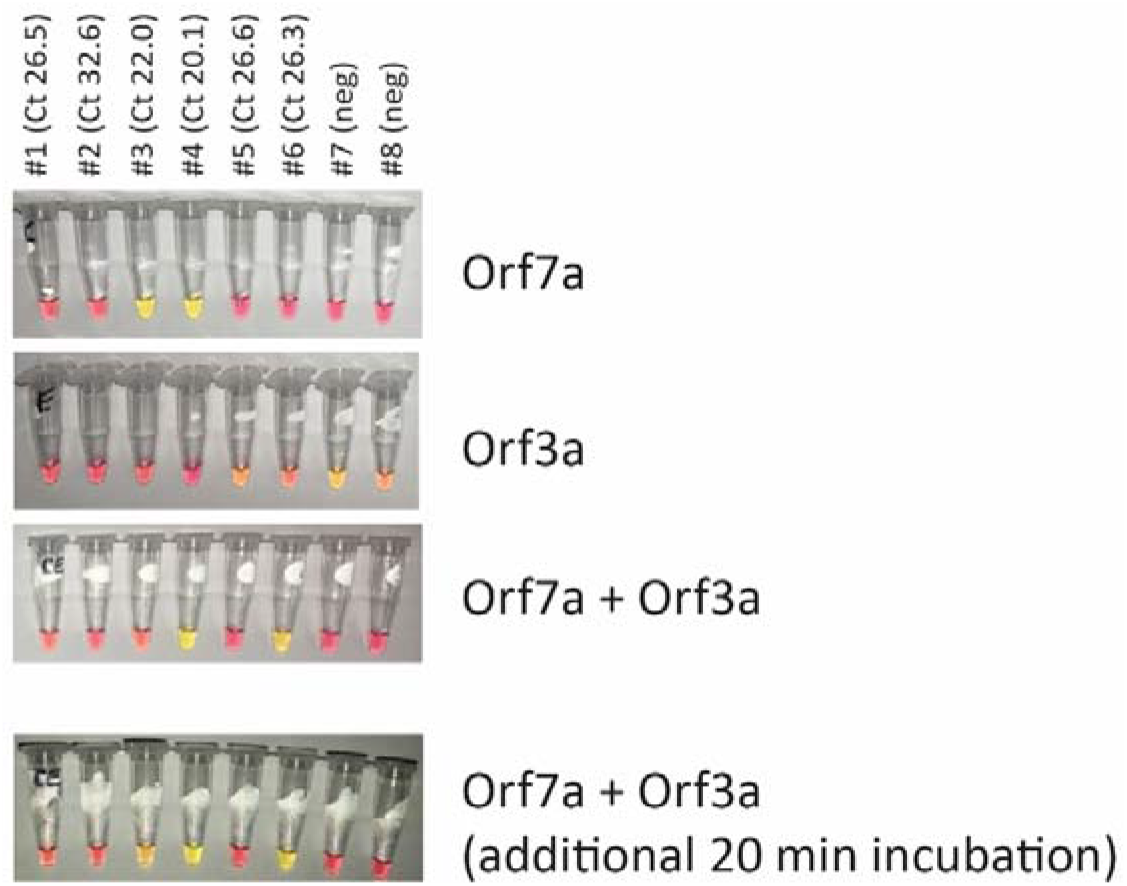
Multiplexing of LAMP reactions. Representative results from experiments trying to combine different LAMP reactions in one tube. A multiplexed reaction targeting both Orf7a and Orf3a with a slight elongation of reaction time appears to be more sensitive and specific.

**Fig. S4:**
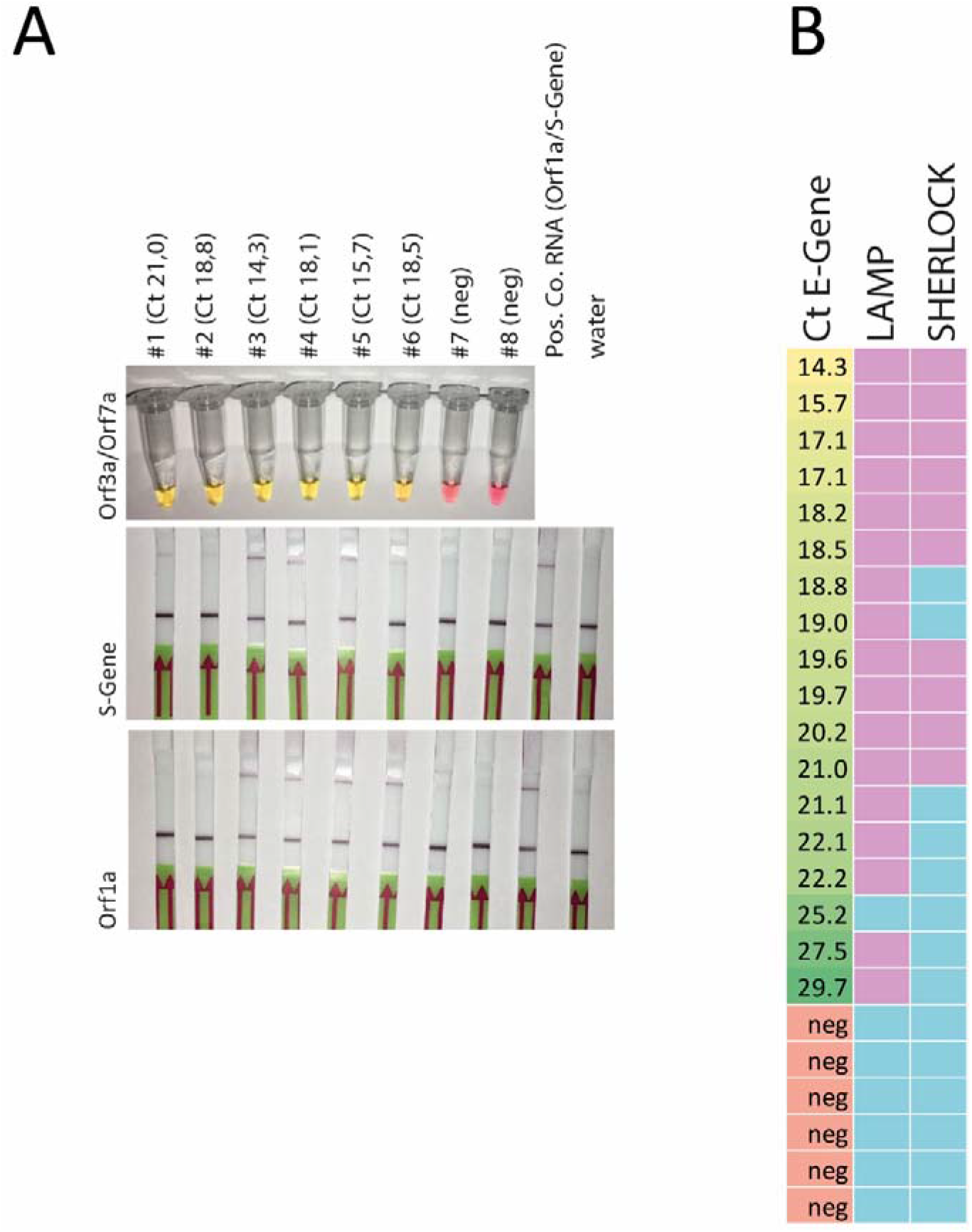
Direct comparison of multiplexed LAMP with two-step SHERLOCK. A. Representative results from 8 samples analyzed by multiplexed LAMP assay (Orf3a and Orf7a) and Sherlock (Orf1a and S gene). B. Summary of all LAMP and Sherlock assays performed in parallel (SHERLOCK assay was regarded as positive with at least one positive result out of two assays; left column: Ct value for E gene from diagnostic PCR in ascending order; green: positive result, blue negative result).

**Suppl. Tab. 1:**
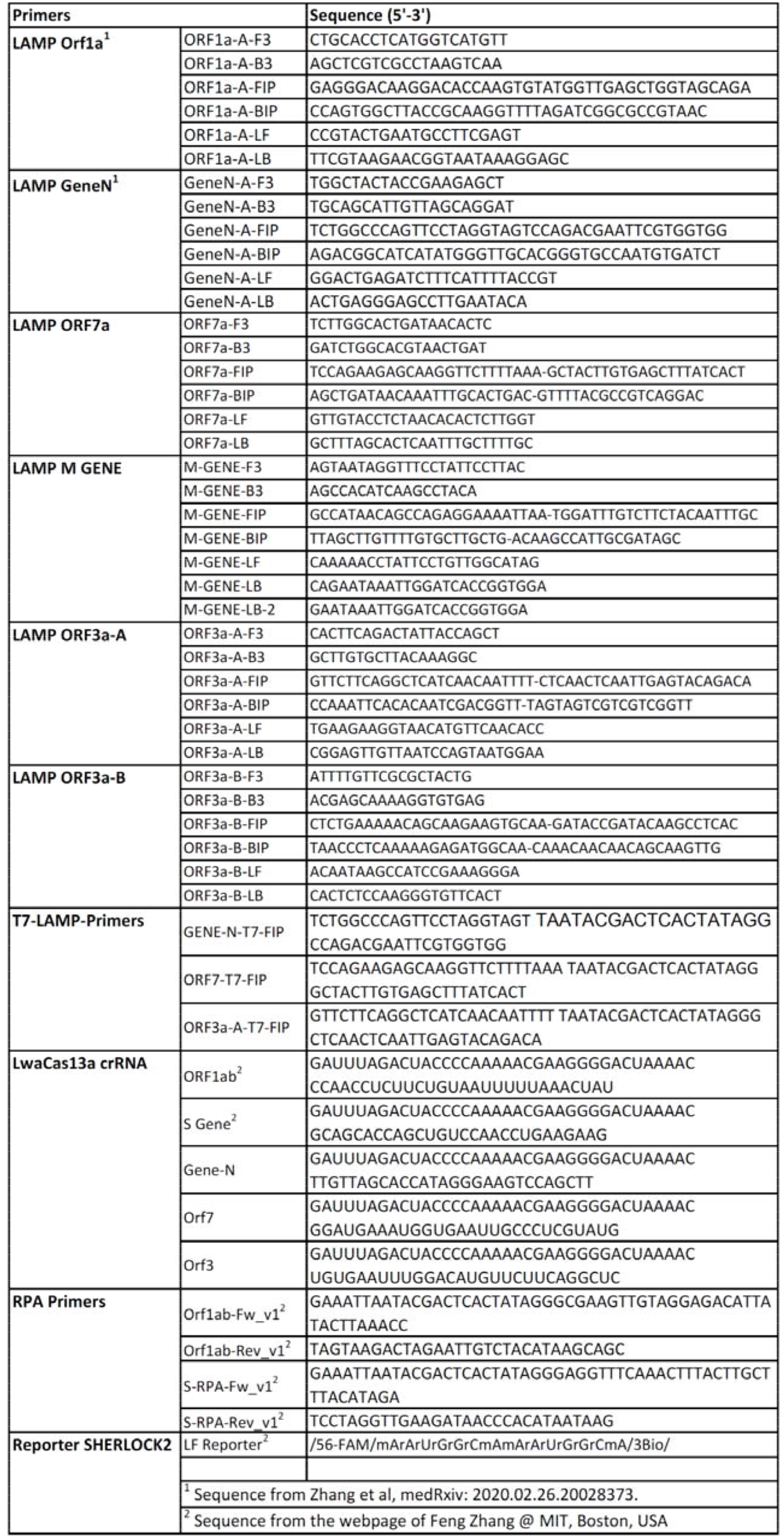
Sequences of all primers and RNAs used in this study

**Suppl. Tab. 2:**
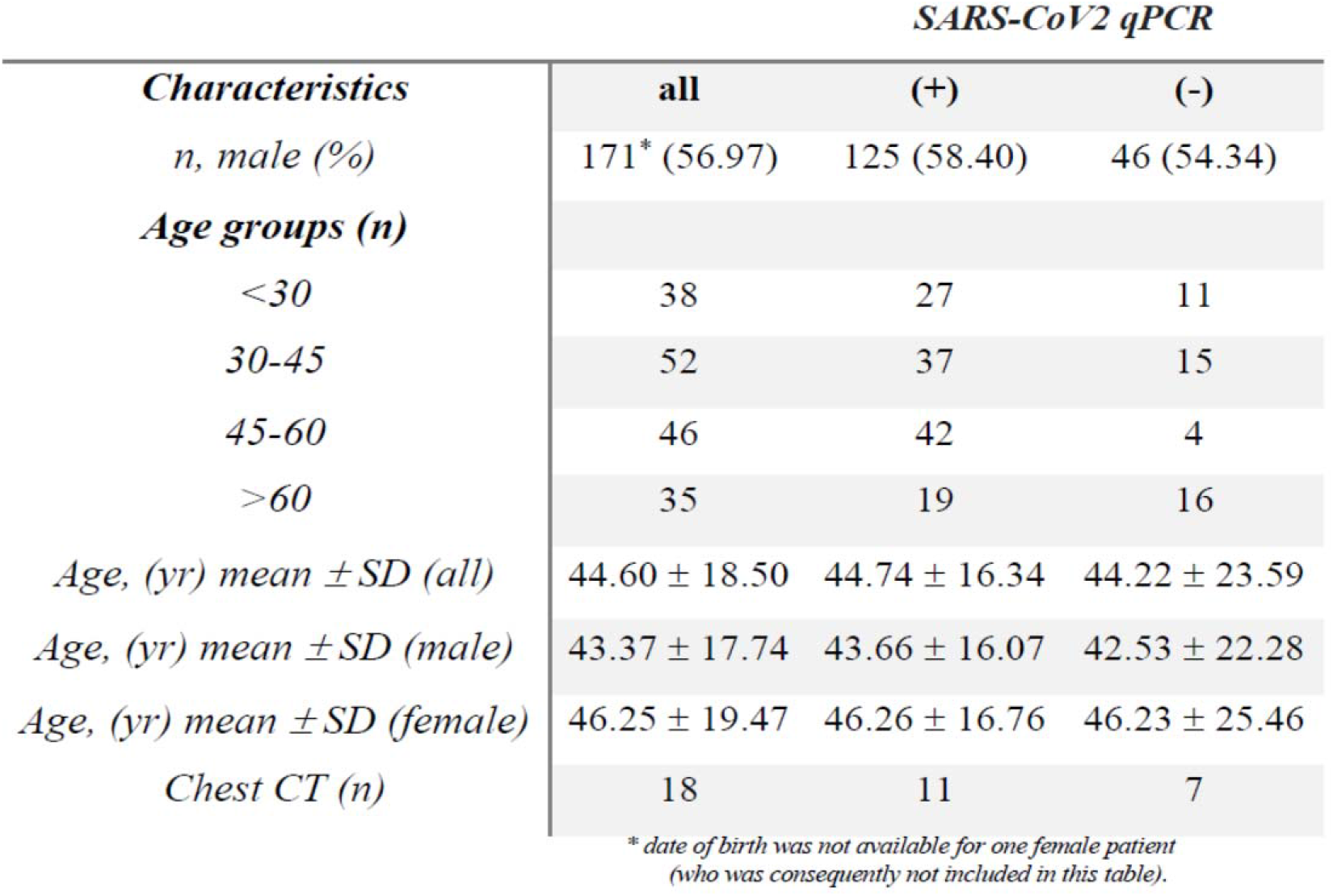
Basic characteristics of the cohort.

